# *RiskPath*: Explainable deep learning for multistep biomedical prediction in longitudinal data

**DOI:** 10.1101/2024.09.19.24313909

**Authors:** Nina de Lacy, Michael Ramshaw, Wai Yin Lam

**Affiliations:** Department of Psychiatry, University of Utah, Salt Lake City, Utah; Scientific Computing Institute, University of Utah, Salt Lake City, Utah

## Abstract

**Highlights:**

- Many diseases are increasingly conceptualized as multifactorial, progressive processes
- Robust prediction of progressive disease courses can advance risk stratification and treatment targeting
- RiskPath provides optimizable timeseries AI to predict progressive disease with longitudinal cohort data
- Enhanced explainability and functionality facilitates risk pathway mapping and compact models

**The Bigger Picture:** Identifying persons at elevated risk for a disease outcome is a key prerequisite for targeting interventions to improve health. Current risk stratification tools for common diseases are aging and achieve only moderate performance. Moreover, many diseases are increasingly recognized to be complex outcomes where individual risk is determined not by a single effect modifier but by time-dependent interactions among many contributory factors over the lifecourse. There is an urgent need to improve individual-level prediction for progressive diseases and understand how multifactorial risks interact over time so that risk stratification and accompanying prevention and intervention strategies can be targeted earlier and more effectively in the disease course.

**Summary:** Many diseases are the end outcomes of multifactorial risks that interact and increment over months or years. Timeseries AI methods have attracted increasing interest given their ability to operate on native timeseries data to predict disease outcomes. Instantiating such models in risk stratification tools has proceeded more slowly, in part limited by factors such as structural complexity, model size and explainability. Here, we present RiskPath, an explainable AI toolbox that offers advanced timeseries methods and additional functionality relevant to risk stratification use cases in classic and emerging longitudinal cohorts. Theoretically-informed optimization is integrated in prediction to specify optimal model topology or explore performance-complexity tradeoffs. Accompanying modules allow the user to map the changing importance of predictors over the disease course, visualize the most important antecedent time epochs contributing to disease risk or remove predictors to construct compact models for clinical applications with minimal performance impact.

## Introduction

Progressive diseases such as cardiovascular, neurodegenerative, metabolic and psychiatric disorders and cancer collectively represent >90% of morbidity, mortality and healthcare costs.^1^ Individual risk for these complex conditions is determined by cumulative interactions among many contributory factors, presenting sequential opportunities for mitigation and prevention. Clinical medicine has long focused on developing risk stratification tools that stratify individual risk for progressive diseases in order to implement preventative strategies, target interventions or help choose among treatments. A risk stratification tool typically consists of a predictive model that explains outcome variance in the condition of interest with additional scoring logic to stratify individuals into risk categories such as high, medium or low. Popular tools are available to clinicians and the general public that stratify individual risk for many progressive, chronic diseases. However, these tools are aging, exhibit only moderate predictive performance and were constructed with inputs from a single data collection interval. For instance, predictive accuracy varies from 0.51-0.83 in standard of care risk calculators in chronic obstructive pulmonary disease, diabetes, colorectal and breast cancer and cardiovascular conditions and positive predictive value (PPV, or precision) and specificity are frequently ≤0.15.^2–18^ Motivated by the desire to improve performance and construct models that better approximate the underlying disease process, interest has grown in using methods specialized for learning from timeseries data to predict progressive disease outcomes using longitudinal data. This is a fertile field offering many opportunities for discovery and development given the availability of both classic and emerging population cohorts collecting rich longitudinal data about their participants. Some (e.g. Nurses Health Study; Framingham Heart Study) collected data over decades. New cohorts continue to be formed such as the All of Us, UK Biobank and Million Veteran Program studies. Such observational cohorts with research foci usually collect more varied information than is typically available in an electronic health record with data collection conducted at wider, discrete time intervals, e.g., annually.

Major algorithmic classes that can model timeseries data at the individual level include classical statistical, state space, Bayesian and dynamic linear techniques, functional data analysis and deep learning, or AI. The latter offers specific advantages in modeling the course of progressive disease: it requires no assumptions about priors or the data structure and can learn from native rather than engineered features. Recent work has utilized timeseries AI to construct predictive models for various conditions such as cardiovascular, renal, neurologic and multi-disease outcomes,^19–31^ though more often using electronic health records, physiologic or clinical data and for diagnostic use cases. This prior work has typically not been explainable at the feature level nor optimized the structural configuration of the AI architectures and has obtained heterogenous performance. Of note, timeseries AI architectures easily participate in overparameterized learning. This refers to the behavior of complex algorithms with more parameters than the number of training data points, which is commonly seen in ‘black box’ models such as deep or ensemble learning. For instance, the parameters of a deep learning model are the total number of weights and biases. In these scenarios, models initially follow the traditional bias-variance curve seen in classical machine learning where test error decreases and then increases. However, if parameters are further increased, test error decreases again to a level below the nadir of the initial bias-variance curve. This phenomenon has been dubbed ‘double descent.’ Thus, despite the ostensible potential for overfitting, overparameterized models perform very well empirically and the mathematical basis for this behavior has recently been explained.^32,33^ Since timeseries AI architectures are structurally complex, they will almost always be overparameterized unless applied in extremely large datasets. Moreover, this property can be a strength that improves performance beyond the bias-variance learning regime by increasing the model’s capacity to learn complex patterns, generalize well and converge to good minima.

Many extant predictive studies using timeseries AI in biomedicine have also used a large and/or unconstrained number of predictors without emphasizing explainability. These characteristics tend to limit their potential for practical deployment within patient-facing risk stratification tools and make them more suited as automated decision-support or diagnostic tools running in high-capacity environments, where it is possible to passively ingest all required inputs from an electronic health record. Creating explainable models can also facilitate engagement with expert clinical opinion to shape and make meaning in predictive modeling. Moreover, in many settings it is desirable to use smaller and simpler models given resource constraints or user preference. Since clinicians and patients have engaged in risk stratification in common progressive diseases for decades, they are accustomed to using tools that ask for ≤10 specific and typically recognizable pieces of data. Finally, the full potential of explainable timeseries AI modeling has not been exploited. Since these algorithms can operate on native longitudinal data, they present the opportunity to inform intervention and prevention discovery by interrogating, for example, how risk interactions evolve over time epochs leading up to the disease outcome.

To address these opportunities, we present RiskPath, a toolbox for timeseries prediction in longitudinal tabular cohort data. RiskPath offers three leading advanced architectures for learning in timeseries data as well as comparisons with popular classical machine learning approaches. Automated optimization of deep learning topology is integrated into predictive modeling to facilitate the production of robust models with optimized architectures. Users can also explore performance-complexity tradeoffs to quantify the performance impact of configuring a simpler model with fewer neurons. Similarly, users may want to reduce the number of predictors in their models given expert input, deployment constraints or to achieve smaller models to improve usability. The feature ablation module visualizes the relative importance of predictors to help guide these decisions. RiskPath enhances translational explainability by embedding the Gradient SHAP^34,35^ process, a game-theoretic approach for gradient-based models, and using its output to generate novel metrics that delineate cumulative risk pathways.

In this study we illustrate RiskPath’s capabilities and performance in three benchmark, ongoing cohort studies that collect the type of rich tabular data historically used in preparing predictive models for instantiation in risk stratification tools. The Adolescent Brain Cognitive Development (ABCD) cohort is the largest long-term study of child and adolescent development in the US. The Multi-Ethnic Study of Atherosclerosis (MESA) is designed to investigate the development and progression of cardiovascular and metabolic disease in a diverse, multi-ethnic population. The Cardiovascular Health Study (CHS) is a large, long-term observational study focused on understanding cardiovascular disease in older adults. Eight different outcomes were predicted across these cohorts that are examples of highly prevalent progressive disease processes: anxiety; depression; attention deficit hyperactivity disorder (ADHD); disruptive behaviors; total mental illness burden; hypertension; borderline hypertension and metabolic syndrome.

We show that RiskPath achieves very robust performance and provides new ways to map and visualize cumulative risk pathways for progressive diseases. Our work also contributes to the evolving literature on overparameterized machine learning and feature ablation in tabular timeseries data, with practical demonstrations in several large, independent real-world cohorts.

## Results

### Overview of the Toolbox

RiskPath offers three advanced architectures for timeseries prediction with three-dimensional (3D) tabular data: Transformer; Temporal Convolutional Network; and Long Short-Term Memory (LSTM) Network. Leading methods treating the tabular timeseries as two-dimensional (2D) data are also available: Feedforward Artificial Neural Network (ANN); Random Forest (RF); Support Vector Machine (SVM) and Logistic Regression (LR) with the elastic net regularization. Users may select one, any or all methods to perform comparisons. An integrated optimization layer for deep learning architectures identifies optimal network width (number of hidden units) in a principled, automated fashion and allows the user to explore performance-complexity tradeoffs across structural configurations. The feature ablation module allows users to examine relationships between feature importance and performance. RiskPath offers new metrics to interrogate fitted 3D timeseries models that utilize the raw output of Gradient SHAP. Predictor Path computes and visualizes how predictor importance changes over time epochs. Epoch Importance computes and visualizes the relative importance of different time epochs in the timeseries. A number of utilities are also packaged with RiskPath including visualization tools, feature selection with the LASSO (linear) and Boruta (nonlinear) methods and 3D matrix concatenation with five different options for padding incomplete timeseries matrices. Here, we demonstrate results after applying RiskPath to predict depression, anxiety, disruptive behaviors, ADHD and total mental illness burden in the ABCD cohort; hypertension and borderline hypertension in the CHS cohort; and metabolic syndrome in the MESA cohort.

### Performance of timeseries and classical machine learning methods are similar

Performance of optimized timeseries AI models and the best performer among optimized feedforward neural networks, Random Forest, SVM and logistic regression models was similar (**Table 1a and 1b**). Among the three timeseries architectures available in RiskPath, temporal convolutional networks and transformers performed better than LSTMs. The margin of improvement is idiosyncratic among learning tasks. For instance, transformers or temporal convolutional networks improve accuracy by 1-2% in mental illness prediction in ABCD, but do 8-10% better in predicting metabolic syndrome in the MESA cohort. To assess the degree of performance sensitivity to sample size, we sub-sampled the data and repeated our analyses in samples of size 50% and 25% of original samples for each experiment. Performance across techniques and predictive targets did not markedly differ in the sensitivity analysis. These results may be inspected in **Supplementary Table 1**. In addition, plots of Receiver Operating Characteristic (ROC) curves are available in **Supplementary Figure 3**.

**Table 1 a and 1b:** Comparative performance of timeseries AI and machine learning methods in predicting eight disease conditions across the ABCD, MESA and CHS cohorts. Performance in out of sample testing is shown for baseline models obtained with all inputs remaining after feature selection (**Supplementary Table 2**). Algorithm settings may be viewed in **Supplementary Table 3**. A Temporal Convolutional Network and a Transformer model are abbreviated as TCN and Trans respectively.

| *a* | <u>Model</u> | <u>Accuracy</u> | <u>AUROC</u> | <u>Precision</u> | <u>Recall</u> | <u>Specificity</u> | <u>F1</u> | <u>Loss</u> | <u>Time (secs)</u> |
| --- | --- | --- | --- | --- | --- | --- | --- | --- | --- |
| **ADHD**<br>*n*=754 | LSTM | 0.951 | 0.989 | 0.953 | 0.951 | 0.953 | 0.951 | 0.475 | 53 |
|  | TCN | 0.966 | 0.992 | 0.967 | 0.966 | 0.967 | 0.966 | 0.140 | 44 |
|  | Trans | 0.966 | 0.992 | 0.966 | 0.966 | 0.966 | 0.966 | 0.096 | 99 |
| **Anxiety**<br>*n*=928 | LSTM | 0.948 | 0.980 | 0.951 | 0.948 | 0.951 | 0.948 | 0.680 | 61 |
|  | TCN | 0.960 | 0.992 | 0.961 | 0.960 | 0.961 | 0.960 | 0.118 | 102 |
|  | Trans | 0.958 | 0.990 | 0.958 | 0.958 | 0.958 | 0.958 | 0.125 | 96 |
| **Depression**<br>*n*=824 | LSTM | 0.932 | 0.981 | 0.936 | 0.932 | 0.936 | 0.932 | 0.602 | 55 |
|  | TCN | 0.955 | 0.990 | 0.955 | 0.955 | 0.955 | 0.955 | 0.136 | 128 |
|  | Trans | 0.946 | 0.989 | 0.947 | 0.946 | 0.947 | 0.946 | 0.144 | 104 |
| **Disruptive**<br>*n*=518 | LSTM | 0.968 | 0.997 | 0.968 | 0.968 | 0.968 | 0.968 | 0.683 | 42 |
|  | TCN | 0.977 | 0.998 | 0.977 | 0.977 | 0.977 | 0.977 | 0.091 | 143 |
|  | Trans | 0.982 | 0.999 | 0.982 | 0.982 | 0.982 | 0.982 | 0.051 | 56 |
| **Total MI**<br>*n*=1266 | LSTM | 0.961 | 0.989 | 0.961 | 0.961 | 0.961 | 0.961 | 0.602 | 78 |
|  | TCN | 0.970 | 0.990 | 0.970 | 0.970 | 0.970 | 0.970 | 0.152 | 154 |
|  | Trans | 0.970 | 0.992 | 0.970 | 0.970 | 0.970 | 0.970 | 0.116 | 151 |
| **Hypertension**<br>*n*=1332 | LSTM | 0.837 | 0.919 | 0.838 | 0.837 | 0.838 | 0.837 | 0.377 | 1865 |
|  | TCN | 0.855 | 0.937 | 0.855 | 0.855 | 0.855 | 0.855 | 0.323 | 831 |
|  | Trans | 0.874 | 0.950 | 0.874 | 0.874 | 0.874 | 0.874 | 0.301 | 2728 |
| **Borderline Hypertension**<br>*n*=418 | LSTM | 0.822 | 0.881 | 0.828 | 0.822 | 0.828 | 0.821 | 0.685 | 723 |
|  | TCN | 0.844 | 0.863 | 0.847 | 0.844 | 0.847 | 0.844 | 0.547 | 455 |
|  | Trans | 0.844 | 0.871 | 0.844 | 0.844 | 0.844 | 0.844 | 0.445 | 497 |
| **Metabolic Syndrome**<br>*n*=3110 | LSTM | 0.808 | 0.896 | 0.809 | 0.808 | 0.809 | 0.808 | 0.448 | 164 |
|  | TCN | 0.888 | 0.961 | 0.888 | 0.888 | 0.888 | 0.888 | 0.253 | 84 |
|  | Trans | 0.907 | 0.970 | 0.907 | 0.907 | 0.907 | 0.907 | 0.225 | 364 |

| *b* | <u>Model</u> | <u>Accuracy</u> | <u>AUROC</u> | <u>Precision</u> | <u>Recall</u> | <u>Specificity</u> | <u>F1</u> | <u>Loss</u> | <u>Time (secs)</u> |
| --- | --- | --- | --- | --- | --- | --- | --- | --- | --- |
| **ADHD**<br>*n*=754 | LR | 0.969 | 0.993 | 0.970 | 0.970 | 0.970 | 0.969 | 0.098 | 6 |
|  | RF | 0.982 | 0.994 | 0.982 | 0.982 | 0.982 | 0.982 | 0.202 | 3 |
|  | SVM | 0.963 | 0.993 | 0.963 | 0.963 | 0.963 | 0.963 | 0.093 | 2 |
|  | ANN | 0.954 | 0.990 | 0.955 | 0.954 | 0.955 | 0.954 | 0.229 | 8 |
| **Anxiety**<br>*n*=928 | LR | 0.953 | 0.991 | 0.953 | 0.953 | 0.953 | 0.953 | 0.121 | 6 |
|  | RF | 0.955 | 0.992 | 0.956 | 0.955 | 0.956 | 0.955 | 0.195 | 3 |
|  | SVM | 0.951 | 0.991 | 0.951 | 0.951 | 0.951 | 0.951 | 0.125 | 3 |
|  | ANN | 0.943 | 0.980 | 0.943 | 0.943 | 0.943 | 0.943 | 0.384 | 9 |
| **Depression**<br>*n*=824 | LR | 0.944 | 0.989 | 0.944 | 0.944 | 0.944 | 0.944 | 0.134 | 6 |
|  | RF | 0.955 | 0.990 | 0.955 | 0.955 | 0.955 | 0.955 | 0.188 | 3 |
|  | SVM | 0.949 | 0.991 | 0.950 | 0.949 | 0.950 | 0.949 | 0.127 | 2 |
|  | ANN | 0.932 | 0.981 | 0.933 | 0.932 | 0.933 | 0.932 | 0.406 | 8 |
| **Disruptive**<br>*n*=518 | LR | 0.986 | 0.999 | 0.986 | 0.986 | 0.986 | 0.986 | 0.055 | 6 |
|  | RF | 0.982 | 0.999 | 0.982 | 0.982 | 0.982 | 0.982 | 0.126 | 3 |
|  | SVM | 0.982 | 0.998 | 0.982 | 0.982 | 0.982 | 0.982 | 0.065 | 3 |
|  | ANN | 0.982 | 0.996 | 0.982 | 0.982 | 0.982 | 0.982 | 0.194 | 8 |
| **Total MI**<br>*n*=1266 | LR | 0.972 | 0.993 | 0.972 | 0.972 | 0.972 | 0.972 | 0.097 | 6 |
|  | RF | 0.968 | 0.994 | 0.968 | 0.968 | 0.968 | 0.968 | 0.167 | 3 |
|  | SVM | 0.973 | 0.993 | 0.973 | 0.973 | 0.973 | 0.973 | 0.095 | 12 |
|  | ANN | 0.966 | 0.989 | 0.967 | 0.966 | 0.967 | 0.966 | 0.216 | 9 |
| **Hypertension**<br>*n*=1332 | LR | 0.853 | 0.928 | 0.854 | 0.853 | 0.854 | 0.853 | 0.383 | 8 |
|  | RF | 0.837 | 0.925 | 0.837 | 0.837 | 0.837 | 0.837 | 0.398 | 4 |
|  | SVM | 0.855 | 0.938 | 0.855 | 0.855 | 0.855 | 0.855 | 0.321 | 20 |
|  | ANN | 0.841 | 0.913 | 0.841 | 0.841 | 0.841 | 0.841 | 0.450 | 113 |
| **Borderline Hypertension**<br>*n*=418 | LR | 0.800 | 0.878 | 0.800 | 0.800 | 0.800 | 0.800 | 0.477 | 6 |
|  | RF | 0.817 | 0.905 | 0.820 | 0.817 | 0.820 | 0.816 | 0.457 | 2 |
|  | SVM | 0.822 | 0.899 | 0.825 | 0.822 | 0.825 | 0.822 | 0.413 | 1 |
|  | ANN | 0.833 | 0.870 | 0.835 | 0.833 | 0.835 | 0.833 | 0.491 | 93 |
| **Metabolic Syndrome**<br>*n*=3110 | LR | 0.895 | 0.966 | 0.895 | 0.895 | 0.895 | 0.895 | 0.238 | 7 |
|  | RF | 0.850 | 0.935 | 0.850 | 0.850 | 0.850 | 0.850 | 0.396 | 4 |
|  | SVM | 0.891 | 0.964 | 0.891 | 0.891 | 0.891 | 0.891 | 0.243 | 54 |
|  | ANN | 0.843 | 0.923 | 0.844 | 0.843 | 0.844 | 0.843 | 0.415 | 12 |

As expected, classical approaches were faster in training models since they do not operate on native timeseries data, have simpler architectures and do not undergo topologic optimization. Training times for deep learning models (including the feedforward network) are in the range of 1 (smaller ABCD samples) to 45 minutes (larger CHS samples). Of note, these training times are for the entire topologic optimization process which here trains 42 models. Therefore, the training time of an individual deep learning model is much less than the total training time reported in **Table 1b**. All optimized test models, which represent the prototypes that would be embedded in risk stratification tools, including timeseries AI, require < 2 seconds to complete.

### Topologic optimization improves performance of deep learning architectures

For deep learning architectures including timeseries AI and feedforward neural networks, we used RiskPath to vary network width (number of neurons or hidden units) in the range [8,1200] and find the optimal structural configuration with best generalization performance in out of sample testing. This point is identified by finding the best test performance in the portion of the learning curve where test accuracy and loss have stabilized. **Figure 1** shows example RiskPath visualizations of how topologic optimization is achieved for predicting anxiety with different timeseries AI architectures. In these performance-complexity curves, performance increasingly improves as network width is increased past the classical bias-variance zone and interpolation point before test accuracy and loss flattens. Red triangles in these plots indicate optimal model topology. This is typical behavior, as may be seen in other disease targets in **Supplementary Figure 1**.

**Figure 1:**
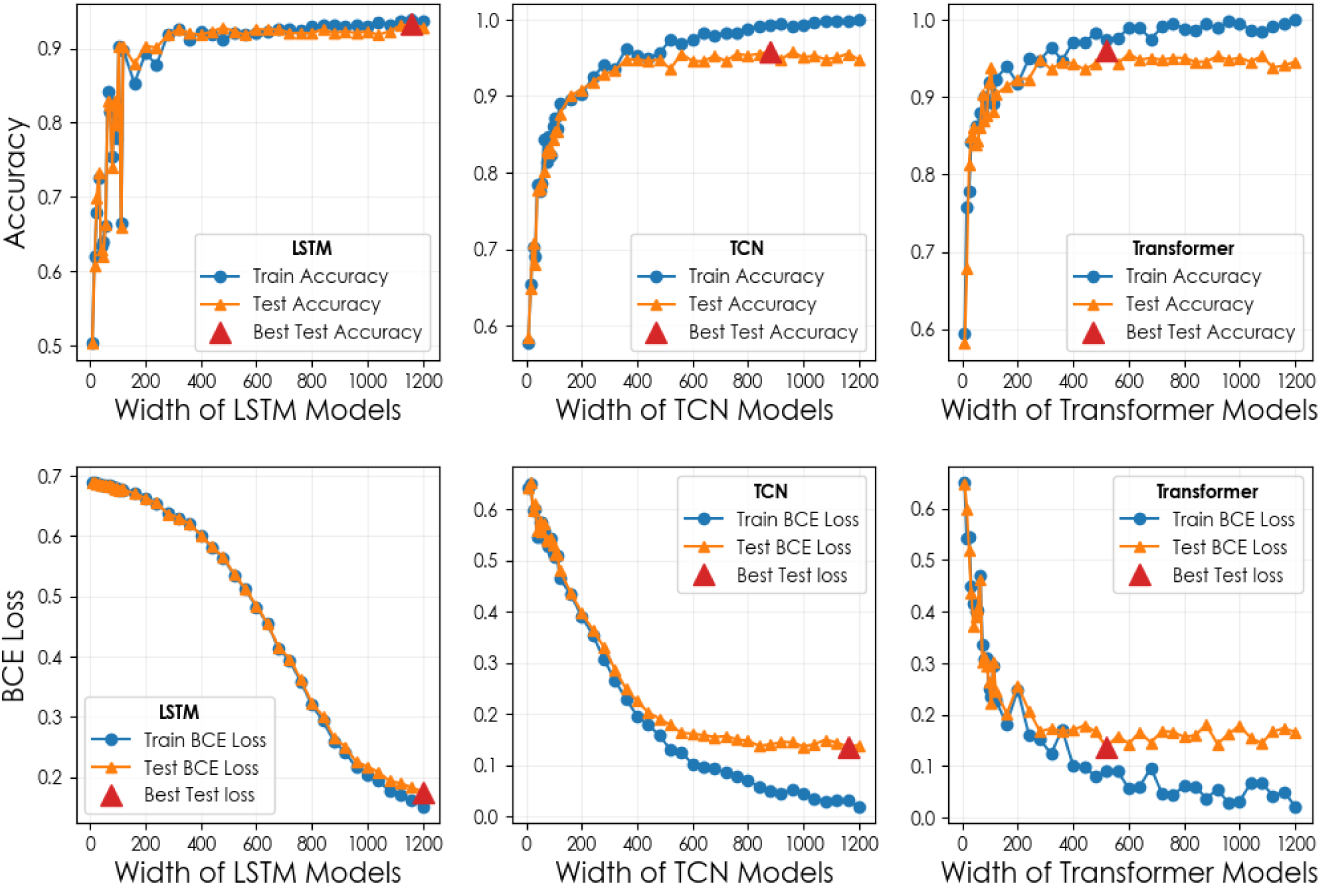
Performance- complexity tradeoffs in prediction of anxiety with three different timeseries AI architectures. The relationship between increasing network width and accuracy (top) or binary cross-entropy loss (bottom) is shown for predicting anxiety with LSTM, temporal convolutional network or transformer architectures in the ABCD cohort. Red triangles indicate the optimal structural configuration identified by RiskPath. A Temporal Convolutional Network is abbreviated as TCN.

### Feature ablation has minimal impact on or improves generalization

Fine tuning fitted predictive models by removing features may be motivated by the desire for simpler or more compact models, data or system constraints or in response to expert input. The feature ablation module in RiskPath provides a principled basis for feature reduction in fitted models. Here, predictor importances calibrated with mean SHAP values are analyzed to identify the point of diminishing returns where additional features are “no longer worth the corresponding performance benefit.”^36^ **Figure 2** gives an example of this process using a Transformer to predict anxiety. Predictor importance may be observed to quickly decrease and asymptote over weaker features, where the latter are excellent candidates for ablation. An obvious ‘knee’ is visible representing the point of diminishing returns. This typical behavior may be seen in other disease prediction models in **Supplementary Figure 2**. RiskPath embeds Lavorini’s kneefinder function^37^ to automatically find this point by determining the maximum distance between the importance curve and a cord from first to last importance (**Figure 2a**). Users can also specify a fixed number of predictors and re-fit more compact models with the most important predictors meeting this threshold. We applied these processes across the best performing timeseries AI models to quantify the impact of feature ablation using the most important features up to the point of diminishing returns and the 10 most important features, where the latter corresponds to the number most often seen in risk stratification tools. This *post hoc* analysis showed that ablating features based on their importances results in minimal performance impact (**Table 2**) or minor performance improvement in certain models.

**Figure 2:**
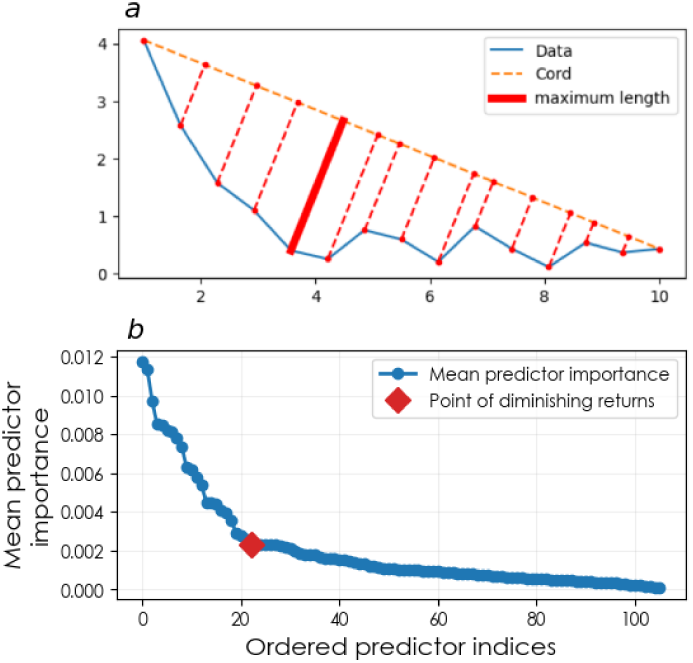
Feature ablation. *a* shows a schematic of how the point of diminishing returns is identified for *b* the prediction of anxiety in the ABCD cohort using a transformer architecture.

**Table 2:** Performance impact of feature ablation in timeseries AI prediction. Performance statistics are shown for the best-performing model in out of sample testing using RiskPath with a) the most important features prior to the point of diminishing returns and b) the 10 most important features. Performance metrics presented correspond to the same individual models as Table 1 after feature ablation.

|  | Before ablation |  | After ablation, point of diminishing returns |  |  |  | After ablation, top 10 features |  |  |  |
| --- | --- | --- | --- | --- | --- | --- | --- | --- | --- | --- |
|  | #Features | Accuracy | # Features | Model | Accuracy | Difference | # Features | Model | Accuracy | Difference |
| ADHD | 106 | 0.966 | 23 | Trans | 0.963 | -0.003 | 10 | TCN | 0.960 | -0.006 |
| Anxiety | 106 | 0.958 | 26 | Trans | 0.948 | -0.010 | 10 | Trans | 0.938 | -0.020 |
| Depression | 88 | 0.946 | 22 | Trans | 0.938 | -0.008 | 10 | TCN | 0.915 | -0.031 |
| Disruptive | 99 | 0.982 | 5 | LSTM | 0.973 | -0.009 | 10 | Trans | 0.986 | 0.004 |
| Total MI | 110 | 0.968 | 27 | Trans | 0.963 | -0.005 | 10 | Trans | 0.959 | -0.009 |
| Hypertension | 106 | 0.862 | 24 | Trans | 0.869 | +0.007 | 10 | Trans | 0.874 | +0.012 |
| Borderline Hypertension | 43 | 0.806 | 9 | Trans | 0.839 | +0.033 | 10 | Trans | 0.828 | +0.022 |
| Metabolic Syndrome | 112 | 0.907 | 16 | Trans | 0.918 | +0.011 | 10 | Trans | 0.914 | +0.007 |

### Novel explainability metrics map risk pathways and epoch contribution to prediction

While SHAP is a popular method for adding explainability to black box models, mean predictor importance across samples is typically the only metric reported, and has not generally been done in timeseries AI studies. A substantial motivation in developing RiskPath to model progressive diseases with timeseries AI is the latter’s ability to operate directly on unmodified tabular data since this opens the opportunity to probe the behavior of predictors over time. RiskPath leverages Gradient SHAP raw output to derive new metrics that allow clinically-relevant interpretation of the timeseries inputs. *Epoch Importance* computes and visualizes the relative importance of different time epochs leading up to disease outcome. An example is shown in **Figure 3** predicting hypertension in the CHS cohort where each time epoch represents a year. Here, it may be observed that the importance of different time epochs to the prediction varies considerably from year to year.

**Figure 3:**
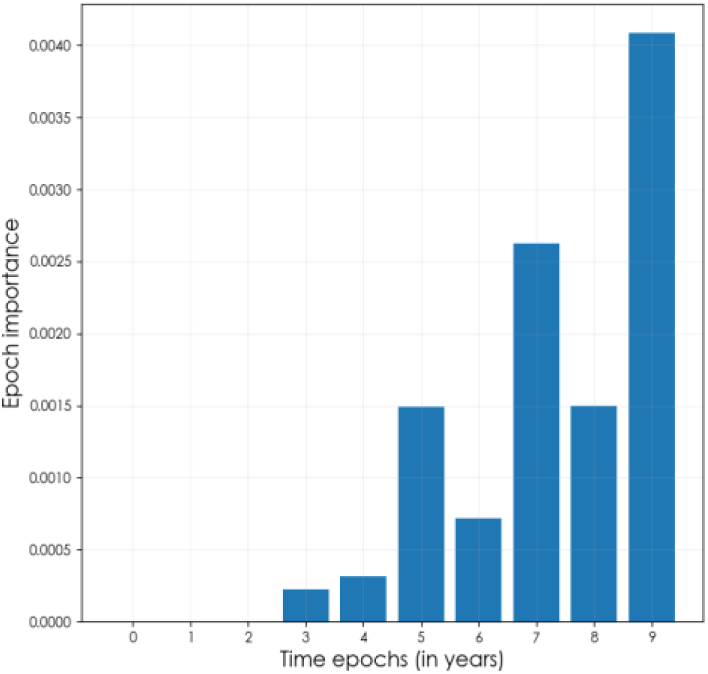
Relative importance of time epochs. The *Epoch Importance* metric is used to compute the relative contribution of each antecedent time epoch risk for hypertension in the CHS cohort.

*Predictor Path* computes and visualizes how predictor importance changes over antecedent time epochs to map the cumulative risk interactions that predict progressive disease outcomes.

The additional granularity offered by this approach shows how individual predictor importance may vary substantially from one period or lifestage to the next. **Figure 4** gives ADHD prediction in the ABCD cohort as an example. In this analysis time epochs correspond to increasing age in children aged 9-13 years. As may be seen, the contribution of screen use time and executive function (cognitive control) deficits to risk for ADHD rises sharply in the run up to adolescence while the importance of other predictors remains relatively stable.

**Figure 4:**
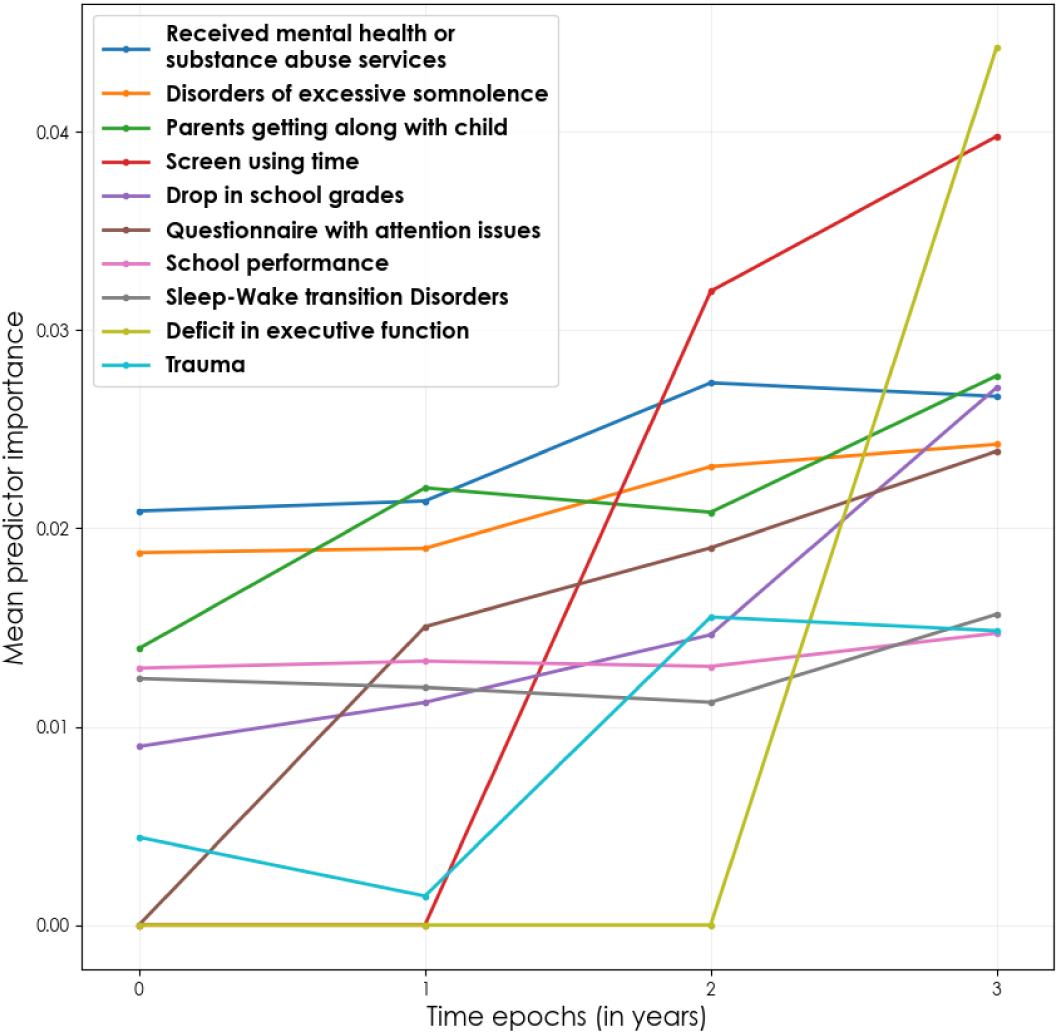
Risk Pathway Mapping. The *Predictor Path* metric is used to compute predictor importance within time epochs and map shifting predictor importance over time.

## Discussion

Interest in using timeseries AI approaches to model disease processes and clinical decision-making has grown in recent years and been further invigorated by the expansion of techniques like transformer architectures.^38^ While there is increasing acceptance of AI for clinical risk stratification, this has been more the case in convolutional neural networks applied to imaging data. These have built on advances in computer vision to produce risk predictions for melanoma,^39^ tuberculosis,^40^ lung cancer,^41^ diabetic retinopathy^42^ and macular edema^43^ with clinically-deployed classifiers that are equal to or better in discriminating cases than board-certified specialists^44–46^ with accuracy of ∼0.88-0.95 and good sensitivity and specificity.^43,46,47^ AI applications using tabular data from longitudinal observational cohorts for predictive modeling, historically among the most classic translational use cases, have gained less traction. Common challenges are the interrelated issues of performance, usability and data collection.^48^ Where a clinical image is a single-shot acquisition of high-dimension data, tabular features in predictive models that are instantiated in risk stratification tools must usually be collected anew and singly from patients. Moreover, iteratively incorporating clinician insights, usability constraints and model optimization promotes the construction of predictive models that generalize well and are parsimonious and user-friendly. In turn, explainability is important to help facilitate these processes.^49^ RiskPath is designed to facilitate easy comparisons and structural optimization for complex timeseries architectures, enhanced explainability and principled approaches to fine-tuning predictors in prediction in longitudinal cohorts.

Our performance comparisons across varied progressive diseases and independent cohorts yielded several interesting observations. Overall, performance in timeseries AI algorithms operating on 3D data and feedforward and classical machine learning techniques operating on 2D data (after Haar wavelet transformation) achieved similar performance. All algorithms achieved 0.85-0.99 across all performance metrics including sensitivity (recall), specificity and PPV, where ≥0.85 is a pragmatic benchmark for clinical utility. This is superior to existing predictive studies using tabular cross-sectional data for risk stratification purposes in mental health (∼0.70-0.80 accuracy and AUROC^50–58^) and cardiovascular and metabolic disease (0.65-0.81 accuracy and AUROC^2–4,59^). Similar performance between ‘3D’ and ‘2D’ models is an expected result, since timeseries AI tends to yield greater performance increments in longer input series than are typically available in a longitudinal cohort.^60^ Notably, timeseries AI pulled ahead a little in the hypertension models, which use data from 10 time epochs. We observed a clear performance separation among timeseries methods with transformer and temporal convolutional architectures excelling. However, there was no single best timeseries architecture, supporting the value of comparing across architectures when fitting a predictive model in a specific dataset. Also as expected, deep learning models took longer to train since these went through the structural optimization process with 42 model fits. The current release of RiskPath was not parallelized in order to provide baseline comparisons with classical techniques. A forthcoming update will include GPU parallelization to speed computations. A simple approach to managing computational overhead in RiskPath would be to survey a smaller and/or sparser range of model widths in the optimization process. Other strategies include model parallelism, training in the cloud, pruning and quantization and checkpointing and resuming. Dropout is also a popular strategy in deep learning, though its application to timeseries learning can be controversial. *Shen et al*. and *Dantas et al* provide further detailed reviews of strategies for improving efficiency in deep learning.^61,62^

We highlight that the above observations are the case *after* optimization of timeseries AI. **Figure 1** shows that alternative structural configurations with smaller model widths to the optimal model selected by RiskPath can yield accuracy and AUROC of 0.75-0.85. This is often – though not exclusively – the performance range observed in existing studies that apply timeseries AI to clinical tabular data (see: **Introduction** for examples), supporting the value of topologic optimization in this algorithmic class. There is no principled method to discover the optimal topology of a deep neural network, and many practitioners use heuristics or trial and error. We approached optimization through the prism of recent theoretical discoveries that explain the empirically strong generalization performance of rich, overparameterized ‘black box’ models that would be expected to overfit under classical bias-variance concepts in machine learning. This phenomenon has been demonstrated in multiple data types and architectures.^63^ We contribute to this literature by showing that an optimization approach that progressively adds network width can take advantage of ‘double descent’ behavior to identify an optimal topologic configuration for deep networks operating on tabular data in eight tasks across 3 independent real-world datasets. This framework can also provide a useful way to quantify the performance impact of choosing less-parameterized deep learning models, such as might be preferred in lower-resource settings or for runtime efficiency. RiskPath visualizations of these performance-complexity tradeoffs can help guide pragmatic decisions regarding these choices. Alternative approaches to altering parameterization include increasing the number of observations (for example by supplementing with synthetic data^64^) or by informing a smaller model with knowledge from a larger model that is less or not overparameterized. Approaches to the latter include transfer learning,^65^ meta-learning,^66^ knowledge distillation^67^ or knowledge graphs.^68,69^ While outside the scope of the current study, additional work in this dynamic field will inform how manipulation of the relationships among structural parameters and the number of input features and observations can optimize learning in timeseries AI.

Model parsimony can be an important issue in translational settings with tabular data collected from patients. In data inputs collected singly, additional assays and assessments can incur additional patient, clinician and administrative burden and accompanying discomfort, pain and/or cost. More generally, parsimony is generally considered to support improved generalization and explainability. The need to transition away from “off-the-shelf” AI and deep learning has been highlighted^70^ and it has been shown that redundancy in feature representations exists even in image or language tasks, permitting ablation and/or recovery training with minimal (or even improved) performance impact.^71,72^ This suggests that the majority of features can be safely ablated without a substantial impact to performance. In the present work, we operationalize these insights by providing a feature ablation module that maps predictor importance to performance metrics and identifies a point of diminishing returns. We tested this across disease conditions and cohorts, finding that the majority of features can be safely ablated to the point of diminishing returns with minimal impact to performance, including sensitivity, specificity and PPV. Further, timeseries AI predictive models for hypertension and metabolic syndrome in the CHS and MESA cohorts showed slightly improved testing performance after ablation in a practical demonstration of parsimony improving generalization. These observations held true even when features were ablated beyond the point of diminishing returns to the tighter threshold of 10 predictors expected by community users of predictive models embedded in risk stratification tools. Taken together, our results suggest that timeseries AI models constructed in tabular, longitudinal cohort data can be safely constrained to much smaller input feature sets with minimal performance impact and in certain datasets even exhibit improved generalization properties.

Enhancing the explainability of AI supports many aspects of model construction. While it is increasingly common for studies using deep learning to report overall or mean predictor importance using explainability techniques like SHAP, this is not typically the case in timeseries AI prediction. Timeseries AI methods offer interesting opportunities to interrogate the developmental course of disease and reason not only about which predictors influence disease risk but also *when* they have the largest impact and *how* cumulative risk pathways shift over lifestages. Here, we construct two new explainability metrics that focus on these latter constructs and complement the conventional mean feature importance metric. Using Epoch Importance, we are able to identify time periods or epochs that make larger contributions to risk for disease outcomes. This could be used to inform prevention or intervention planning. For instance, **Figure 3** suggests that 2 and 4 years prior to the index epoch are relatively more important to risk for hypertension. *Predictor Path* helps visualize how risk predictors shift in importance to inform our understanding of how cumulative disease pathways operate over time. In the example given in **Figure 4**, mapping the cumulative risk interactions that predict ADHD suggests that youth screen time use and executive function, both of which are susceptible to intervention, become increasingly important risk contributors over adolescence.

Taken together, our results across eight different conditions in three large, independent cohorts suggest that timeseries AI approaches that include topologic optimization and enhanced explainability can predict outcomes with parsimonious models that generalize well and inform our understanding of the cumulative risk processes that underpin progressive diseases. While we here apply approaches to classic longitudinal cohorts for the purpose of constructing predictive models angled toward risk stratification in progressive disease, RiskPath may be useful in other biomedical use cases where longitudinal data is available and enhanced translational explainability is valuable such as developmental processes or public health monitoring.

## Experimental Procedures

### Resource Availability

#### Lead contact

Further information and requests will be fulfilled by the lead contact, Nina de Lacy

#### Data and code availability

ABCD data used in this study may be obtained by applying to the ABCD Repository at the National Institute of Mental Health Data Archive (https://nda.nih.gov/abcd): DOI: 10.15154/z563-zd24

CHS data used in this study may be obtained by applying to the BioLINCC repository at the National Institutes of Health (https://biolincc.nhlbi.nih.gov/home/): Accession Number: HLB00040019a

MESA data user in this study may be obtained by applying to the BioLINCC repository at the National Institutes of Health (https://biolincc.nhlbi.nih.gov/home/): Accession Number: HLB00640824a

RiskPath is an open source code repository in Python and PyTorch that is available at our GitHub repository (https://github.com/delacylab/RiskPath).

### Data used in this study

This study has been deemed not human subjects research by the University of Utah Institutional Review Board.

#### ABCD cohort data

The ABCD data used in this study comes from the ABCD open science repository. ABCD is an epidemiologically-informed study launched in 2017 which is collecting data over 10 years from a 21-site cohort of adolescents across the US. Participants (52% male; 48% female) were enrolled at age 9-10 and are currently 13-14 years old. This is a naturalistic, unstratified cohort. Further descriptions of the overall design as well as recruitment procedures and the participant sample may be found in Jernigan et al; Garavan et al; and Volkow et al.^73–75^ and the study website at abcdstudy.org. ABCD collects rich multimodal data of youth participants and their parents. Here, we utilize variables from assessments of physical and mental health, substance use, neurocognition, school performance and quality, culture, community and environment contributed by youth and their parents as well as biospecimens (e.g. pubertal hormone levels) and environmental toxin exposure.^76,77^

ABCD is a longitudinal study where the cohort contains ∼800 twin pairs and non-twin siblings may be enrolled. General inclusion criteria for the present study were a) participants enrolled in the study at baseline (9-10 yrs) who were still enrolled in the ABCD study through 13-14 yrs (*n*=10,093) who were b) youth participants unrelated to any other youth participant in the study (*n*=8,363). If a youth had sibling(s) present in the cohort, we selected the oldest sibling for inclusion.

The baseline feature set comprised the majority of phenotypic variables available from the ABCD study, including data collection site, a proxy for geographic location. Predictive targets were formed from participant scores in the ASEBA Child Behavior Checklist (CBCL), a standardized assessment of mental health in widespread clinical and research use.^78^ Parents rate their child on a 0-1-2 scale on 118 specific problem items which are then used to form continuous subscale scores in clinical dimensions of interest such as Anxiety or Depression. To form binary classification targets, we thresholded and discretized CBCL subscale T scores using cutpoints established by ASEBA for clinical practice by deeming every individual with a T score ≥65 as a positive case (encoded as 1) and every individual with a score <65 as a negative case (encoded as 0) for ADHD (Attention disorder in ASEBA), Anxiety, and Depression, but a T score of ≥60 as the cutpoint for Total Burden of MI. Disruptive Behavior cases are established by meeting these case criteria for either Aggression or Rulebreaking, corresponding to the clinical definition of Disruptive Behavior Disorder. The ABCD data dictionary may be viewed at: https://nda.nih.gov/data_dictionary.html?source=ABCDALL.

#### CHS cohort data

The CHS data that we used in this study comes from NIH’s BioLINCC open science repository. The CHS study (https://chs-nhlbi.org/) was launched in 1987 to identify risk factors for cardiovascular disease related to coronary heart disease in adults aged 65 or older and the investigation of pulmonary disorders, diabetes, kidney disease, vascular dementia, and frailty. CHS collected data annually from 1989-1999 via extensive annual clinical examinations. Measurements included traditional risk factors such as blood pressure and lipids as well as measures of subclinical disease, including echocardiography of the heart, carotid ultrasound, and cranial magnetic-resonance imaging (MRI). This is an observational cohort. Further descriptions of the overall design as well as recruitment procedures and the participant sample may be found in Fried et al.^79^ Participants were contacted by phone to ascertain their health status every 6 months. Clinical outcomes were recorded in a binary fashion and included coronary heart disease, angina, heart failure, stroke, transient ischemic attack, claudication, diabetes and hypertension. Here, we utilize variables collected from patients spanning laboratory assays (e.g. thyroid hormone, glucose and lipid levels), health, medication and mental health history, behavioral, diet, exercise and health habits and clinical testing (e.g. blood pressure, heart auscultation, brain imaging). The CHS data dictionary may be viewed at: https://biolincc.nhlbi.nih.gov/media/studies/chs/data_dictionary/CHS_v2019a.pdf.

Inclusion criteria for the present study are participants who were enrolled (and agreed to share their data) and remained alive in the CHS study for 10 years. This comprised a total of 3,206 participants. The predictive targets in CHS are supplied in the original data as a clinical ternary variable where original data had 1=Normal; 2=Borderline; 3=Hypertension. The present study defined two binary targets accordingly: Hypertension (n=2382) is the comparison between Normal and Hypertension, and Borderline Hypertension (n=1729) is the comparison between Borderline Hypertension and Hypertension.

#### MESA cohort data

The MESA data used in this study come from the NIH’s BioLINCC open science repository. MESA is a longitudinal study aimed at understanding the development of cardiovascular disease across diverse populations. It includes data from over 6,800 participants aged 45-84, with no prior history of cardiovascular disease, representing multiple ethnic groups. The dataset features extensive imaging, genetic, and clinical data. While the dataset contains 6 exams from 2000 to 2018, only the first 4 exams (2000-2007) are considered in this study due to the substantial reduction in sample size in the latter part of the study.

Inclusion criteria of the MESA data in the present study are participants not excluded from all follow-up events (n=6809). The predictive target in MESA is metabolic syndrome where the original data supplied it as a binary variable. By subsetting to the participants who have been assessed with metabolic syndrome in the last considered examination, a total of 5,636 participants are considered in this study. The objectives and design of the study can be found in Bild et al.^80^ The MESA data dictionary may be viewed at https://biolincc.nhlbi.nih.gov/media/studies/mesa/data_dictionary/MESA_v2023a.pdf.

### Data cleaning and preprocessing

For each studied dataset, the study data dictionary guided the variable encoding process. To ensure parsibility as a numerical dataset, all non-numerical variables were removed from consideration. For each variable, values represented as missingness in the dictionary were encoded as missing data points. Phenotypic variables in ABCD lacking summary scores were scaled (by min-max normalization) and averaged to a summary metric. Pairs of variables with one encoding unit of measurements were paired up to ensure interpretability (e.g., ‘How often do you use fat in cooking’ and ‘Unit of time’). The names of variables that changed over time epochs have been standardized to ensure consistency throughout the timeseries dataset.

The pipeline for data preprocessing is summarized in **Figure 5**. All variables after cleaning were encoded according to the dictionary as a nominal, ordinal, or continuous variable. Non-binary nominal variables were one-hot encoded. Variables that could potentially leak information to each predictive target were removed after inspection by NdL, who is a medical doctor. For instance, symptoms of the predictive target in ABCD or medication given for the predictive target in CHS and MESA. Features with >35% missing values were discarded, where prior research shows that good results may be obtained with machine learning methods for imputation with up to 50% missing data.^81^ The resultant data were randomly partitioned into a training and validation partition and a test partition held out for out of sample testing in the ratio 7:3 by stratifying the target. All subsequent pre-processing was performed separately in these partitions to avoid bias and information leakage.

**Figure 5:**
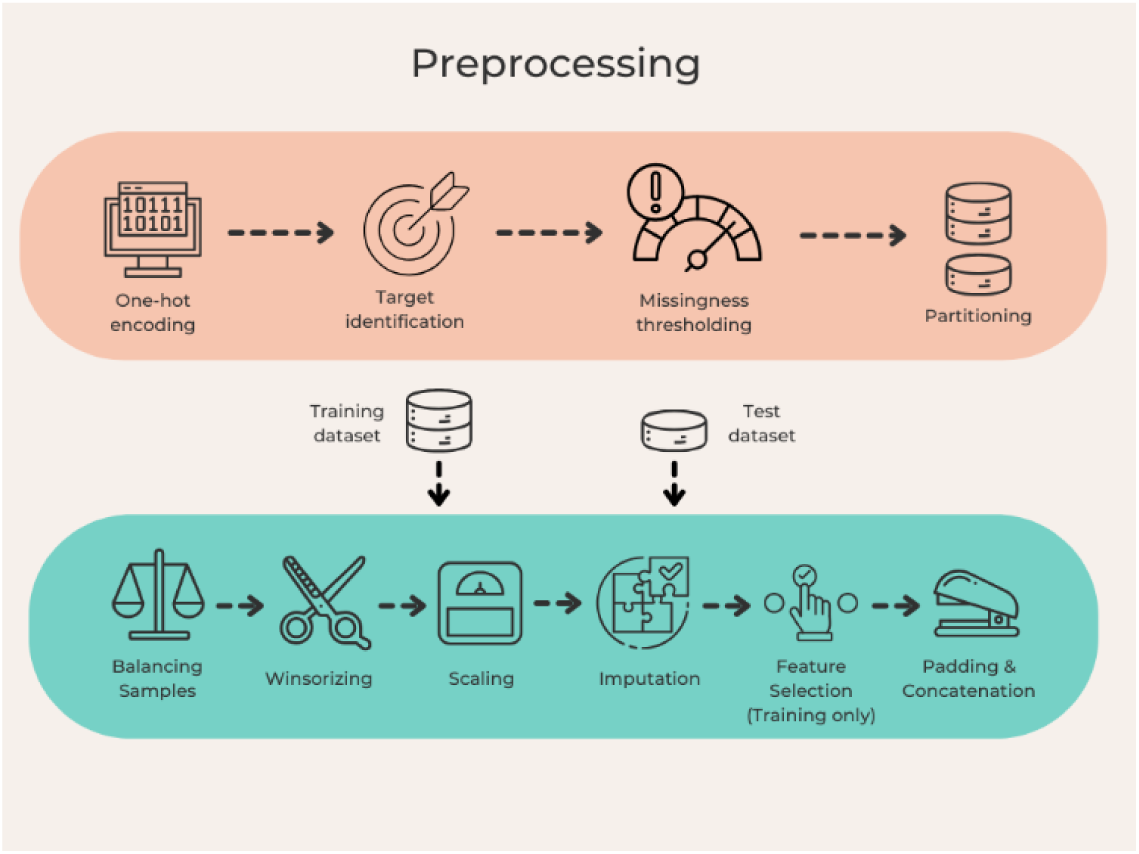
Preprocessing pipeline.

Each subsequent preprocessing step is then applied to the training and test datasets independently within each time epoch. For each positive case of the target, a negative case of the same gender and age group (binarized by median age) was matched. See **Table 3** for sample sizes after balancing.

**Table 3:** Study sample and feature set sizes after feature selection.

|  | #Time epochs | Sample size (n) |  | #Features |  |  |  |
| --- | --- | --- | --- | --- | --- | --- | --- |
|  |  | Training | Test | All | Linear | Nonlinear | Both |
| ADHD | 4 | 754 | 326 | 1035 | 168 | 106 | 225 |
| Anxiety | 4 | 928 | 404 | 1035 | 175 | 106 | 229 |
| Depression | 4 | 824 | 354 | 1035 | 146 | 88 | 197 |
| Disruptive | 4 | 518 | 218 | 1035 | 266 | 99 | 313 |
| Total MI | 4 | 1266 | 562 | 1035 | 204 | 110 | 250 |
| Hypertension | 10 | 1332 | 572 | 1518 | 452 | 106 | 505 |
| Borderline Hypertension | 10 | 418 | 180 | 1510 | 181 | 43 | 207 |
| Metabolic Syndrome | 4 | 3110 | 1334 | 993 | 241 | 112 | 302 |

Continuous features were winsorized to the bounds of mean +/− 3 standard deviations whereas ordinal features were winsorized according to the bounds encoded in the provided data dictionary. Then all features were scaled to the unit interval using min-max normalization. Missing values were imputed using non-negative matrix factorization (with an initial strategy of filling in mean values), an imputation method that is particularly suitable for large-scale multimodal data since it performs well regardless of the underlying pattern of missingness.^82–84^

### Input matrix padding

In longitudinal data, padding may be required to impute features with no samples in a given time epoch. RiskPath offers matrix padding as a utility where users may select among five different padding strategies: *constant*, *mean* (average of samples from other time epochs), *median* (*median* of samples from other time epochs), *backward* (samples from the closest earlier time epoch), and *forward* (samples from the closest later time epoch). Notice that an optimal choice of padding strategy is data-dependent. For the sake of simplicity, each feature without any sample in a time epoch is padded with the constant approach (i.e., padding with 0) in this study.

### Feature Selection

As a utility for user convenience, RiskPath provides 3 options for feature selection: (1) L1-regularized and cross-validated logistic regression to capture features linearly relevant to the target (*linear*), (2) the random-forest-based Boruta^85^ method to capture features non-linearly relevant to the target (*nonlinear*), and (3) combining the feature subsets returned by (1) and (2) (*both*). Users can also choose to opt out of any feature selection technique (*none*) and use their own preferred approach or no feature selection.

(1) is implemented by the *scikit-learn* LogisticRegressionCV^86^ and (2) by the BorutaPy package^87^. RiskPath uses Python wrappers to adapt these methods for time series data. Users have explicit control of settings that constrain the number of features selected by each technique. In the present study, feature subsets of each setting in linear, nonlinear, both, and none were generated for each experiment, and performance was compared across these feature subsets.

The L1-regularized and cross-validated logistic regression approach, also known as the *LASSO CV* in the regression case, is a gold standard linear feature selection technique for ML that identifies a set of linear coefficients minimizing the logarithmic loss of the prediction under L1-regularization. Default settings in LogisticRegressionCV were adopted in our experiments. RiskPath adapted this subroutine for time-series data by separately performing feature selection at each time epoch in the training dataset. The union of features across time epochs was obtained to determine an overall linear feature subset. RiskPath users are offered the additional option of thresholding coefficients output to further constrain the size of feature sets. Our baseline analyses adopted a lenient approach by leaving coefficients un-thresholded so that all features with a non-zero coefficient were accepted as linearly relevant.

Boruta is an ensemble-based technique that is a popular nonlinear method for feature selection. Constructed around a random forest algorithm, it compares original features with their *shadow copies* (original features with values shuffled) such that a feature is important only when it has an importance score higher than the maximal importance score in the set of shadow features, that is, if it has a significantly higher importance over *k* random forest runs than the expected value *k/2* (as defined by a binomial distribution with p = 1/2). Significance is measured by multiple hypothesis testing. Similar to the discussion of the linear subroutine, the Boruta subroutine in RiskPath is separately performed at each time epoch to generate time-specific feature subsets, and then the union of features across epochs is obtained to determine an overall nonlinear feature subset for the experiment. RiskPath offers users the ability to control two tunable hyperparameters that control the strictness of the selection criteria. The first is the level of significance alpha that can be adjusted to expand or shrink the rejection region in each two-sided hypothesis test and concomitantly the size of the feature subset. The second is the percentile (perc) where each real feature is compared to a specific percentile of the importance scores of the shadow feature set (instead of the maximal case when perc=100). Intuitively, setting a higher alpha or lower perc will return a larger number of important features at a higher risk of false positives. In the present study, we implemented Boruta with alpha=0.05 and perc=100, and each embedded random forest with a tree depth of 7 to avoid runtime overhead. **Table 3** shows the number of features after feature selection and **Supplementary Table 2** shows the feature sets that were used in each experiment after the feature selection using each method. In the present study, results reported for the best-performing models are those exhibiting best performance out of linear, nonlinear or linear+nonlinear feature selection.

### Timeseries AI architectures and settings

LSTMs incorporates 3 gates to regulate the flow of information: the forget gate (discarding old information), the input gate (storing new information), and output gate (controlling output at each time epoch). Commonly, practitioners use bidirectional layers to direct a LSTM model forward and backward in timeseries or sequence to capture a richer context representation. Generically, the number of parameters in an LSTM is 4(*hm + h2* + *h*) where *h* = number of hidden units and *m* = the number of features, and a bidirectional LSTM doubles that of a unidirectional LSTM. LSTM models in RiskPath are trained with two bidirectional layers with the tanh activation function with its result in the last time epoch for probability estimation.

Temporal Convolutional Network is a parallelized process that considers the entire timeseries at once by utilizing convolutional layers and dilation factors to handle sequential data. Temporal Convolutional Networks consist of three factors that determine the size of the model parameters: kernel size *k* (size of the filter applied in the convolution), input channels *C_in_* (number of multivariate sequences), and output channels *C_out_* (number of filters in the layer). The number of trainable parameters in a Temporal Convolutional Network is represented by the sum of *k × C_in,i_ × C_out,i_* + *C_out,i_* over each convolutional layer *i* (where *C_in,1_* = the number of features). Temporal Convolutional Networks are composed of individual temporal blocks, each consists of 2 one-dimensional convolutional layers (activated by a ReLU function) to connect the input and output channels. The convolutional layer is equipped with a kernel size *k* of 3, a doubling size of dilation *d* over subsequent blocks (starting at 1), a stride of 1, and a padding size of *d*(*k* - 1)/2. Another pair of layers (with a kernel size of 1) is applied for the first temporal block when the numbers of input and output channels mismatch. Each Temporal Convolutional Network model in RiskPath is sequentially composed of two temporal blocks and lastly a fully connected 1-unit layer for probability estimation.

Transformers encode features into a higher dimensional space where a self-attention mechanism allows transformers to focus on relevant time epochs automatically. The model embeds each of the *m* features to a higher dimensional space of size *d* yielding (*m*+1) *× d* parameters, encode the timeseries (of length *t*) by *positional encoding* with *td* parameters. The attention phase consists of a 3-step projection (i.e., query, key, and value) and an output projection, yielding 4*d*^2^ parameters, then passes to a (usually 2-layer) feedforward network with a width *w,* yielding 2*dw* + 3*d + w* parameters after normalization. The total number of parameters in a *k*-layer transformer model is simplified as *d*(*t* + *m*+1) + *k*(4*d*^2^ + 2*dw* + 3*d* + *w*). Transformer models begin with projecting the features to the tunable embedding space, position the sequential encoding by the number of time epochs, carry forward to a two-layer encoder with 1024 hidden units and 8 attention heads, and bridge to a fully connected 1-unit layer for probability estimation.

For all deep learning models (timeseries and feedforward, below), the hidden weights of all mentioned algorithms are initialized with the Xavier uniform distribution (where LSTM models have an orthogonal distribution for their hidden-to-hidden weights), and biases are initialized as 0. Binary cross-entropy loss is selected as the loss function used for model fitting. In the present study, the AdamW optimizer is used with weight decay = 0.1 and learning rate = 10^−5^. Models were trained for a maximum of 150 epochs with early stopping (patience = 5, evaluated by validation loss). RiskPath allows these settings to be modified by the practitioner for local heuristics or experimental preferences. All embedded deep-learning algorithms and the GradientSHAP values computation in RiskPath are encoded with PyTorch^88^ and Captum^89^ packages in Python respectively.

### Comparison machine learning techniques and settings

Standard feedforward artificial neural network and three classic machine learning approaches are available in RiskPath and investigated in the present study: feedforward artificial neural networks, logistic regression, random forest, and SVM. Since these algorithms are not designed for time-series prediction, a Haar wavelet transformation (with decomposition level=1) is applied to the timeseries to obtain a 2-dimensional data matrix that adequately capture the temporal and frequency information of the timeseries data. This is a typical procedure when using these algorithms in time series data. The wavelet transformation is implemented with the PyWavelets package in Python.^90^

Each feedforward model was constructed with 3 hidden fully connected layers with the ReLU activation function equipped to capture potential non-linear relationship between the feature set and the target. By setting each hidden layer with an equal number of units *k*, the number of parameters for a feedforward model is simplified to *k*(*m* + 1) + 2(*k*^2^ + *k*) + *k* + 1 where *m* denotes the number of features. Logistic regression is a learning algorithm designed specifically for classification tasks by modeling the probability of a class label using a logistic function applied to a linear combination of the features. Random Forest is an ensemble-based algorithm that constructs multiple decision tress during training and combines their respective predictions through a voting mechanism to improve accuracy and reduce overfitting. SVM aims to identify the optimal hyperplane to separate data points into classes by maximizing the margin between the closest data points of different classes. These algorithms were encoded with the Python scikit-learn package^86^ and implemented with their default runtime parameters, which are commonly used in benchmarking practices.

### Training and testing procedures

A 10-fold cross-validation technique is incorporated in the model-fitting process. Across the 10 fitted models, the one with the best validated AUROC is used for subsequent evaluation in out of sample testing in the 30% held out data after initial data partitioning. Users can choose from a wide range of metrics (e.g., accuracy, binary cross-entropy, precision, Bayesian information criterion, etc.) to evaluate models. In each experiment, the best-performing model in terms of accuracy (evaluated in out of sample testing) was selected for reporting.

All models were trained and evaluated on High Performance Computing (HPC) clusters, with each node equipped with an AMD EPYC 7713 64-core CPU, 1TB of RAM, and 8 NVIDIA A30 GPUs, each with 24GB of memory. Parallelization was not used for deep learning in the present study.

### Topological optimization process for deep learning models

A common measure of model topology in deep-learning literature is the *width* of the model, referring to the maximal number of units in a hidden layer. In the deep-learning algorithms used in this study and implemented in RiskPath, the width corresponds directly to the number of hidden units in LSTMs and feedforward neural networks, the number of output channels in temporal convolutional networks, and the dimension of the embedding space in transformers. RiskPath adopts a grid search approach by embedding the option of an increasing range of model widths which can be controlled by the practitioner, allowing for the identification of an optimal width value to explore complexity-performance tradeoffs. The suggested setting in our experiments is a grid in [8, 1200], with an interval of 8 in [8, 120) and an interval of 40 in [120, 1200], yielding a total of 42 width values. The small-interval values aim to capture the traditional bias-variance tradeoff zone, with wider intervals for the overparameterized region. This enables practitioners to identify the optimal network width recommended by Risk Path and/or analyze and visualize performance-complexity tradeoffs.

### Feature ablation

Feature ablation is performed by focusing on an important feature subset according to the mean predictor importance of an optimal model in RiskPath. By inputting the raw GradientSHAP values, RiskPath computes the ordered mean predictor importance and automatically identifies the point of diminishing returns (also known as the knee) using the triangle method (**Figure 2**).

### Explainability

Different approximation techniques of Shapley values have been studied as SHAP (SHapley Additive exPlanations). This study utilizes *GradientSHAP* (implemented in Python Captum package^89^), a combined technique of *Integrated Gradients*^91^ and *SmoothGrad*^92^ to compute expected gradients, as a fast approximation of Shapley values for gradient-based models. In the discussion of non-timeseries datasets where the raw SHAP value matrix is 2D, researchers commonly compute the *Mean-Absolute* SHAP values across samples to increase explainability in black box models.^93^ Intuitively, these values capture the relative magnitude of feature importance. However, the same averaging technique is not common in the literature on timeseries analysis. RiskPath explores 3 averaging techniques over the 3D SHAP value matrix to account for different senses of relative importance. To our knowledge, these averaging techniques have not been studied in the relevant literature on time-series analysis.

*Predictor path* marginalizes the distribution of the samples in the 3D SHAP value matrix by computing the absolute values averaged over samples. *Mean predictor importance* further marginalizes the time epoch distribution to encapsulate the general magnitude of feature importance in a timeseries setting. This is performed by computing the absolute values averaged over samples and time epochs. This is further used in the feature ablation module. *Epoch importance* marginalizes the sample and feature distributions as a metric to compare the importance across time epochs. This is performed by computing the absolute values averaged over samples and features.

### Visualization Utilities

RiskPath provides visualization utilities to support predictive analytics in tabular timeseries data. Examples of performance-complexity tradeoff curves (**Figure 1**), mean predictor importance and its point of diminishing returns (**Figure 2**), epoch importance (**Figure 3**), and mean predictor path (**Figure 4**) are shown above. Other visualization tools include Receiver Operating Characteristic (ROC) plots (**Supplementary Figure 3**), stacked bar plots of mean SHAP values (**Supplementary Figure 4**), beeswarm plots that compare feature values and their importances (**Supplementary Figure 5**), and heatmaps for SHAP values across samples and time epochs of a given feature (**Supplementary Figure 6**). RiskPath also provides an animated 3D surface plot for SHAP values to illustrate the pathway of changes in feature importance. Users may consult the GitHub repository of the RiskPath Python package for the tutorial on the full visualization utilities.

### RiskPath Software Package

The RiskPath toolbox is contained in our Python Package available at https://github.com/delacylab/PathLearn/tree/main/RiskPath. The repository provides instructions to install the package, and multiple tutorials explaining how RiskPath models can be created, trained, and evaluated with simple commands including demonstrations in toy datasets, vignettes and example workflows.

## Data Availability

All data produced in the present study are available upon reasonable request to the authors

https://biolincc.nhlbi.nih.gov/home/

https://nda.nih.gov/abcd

## Acknowledgements

This work was supported by the National Institute of Mental Health under award R00MH118359 to NdL.

ABCD data used in the preparation of this article were obtained from the Adolescent Brain Cognitive Development^SM^ (ABCD) Study (https://abcdstudy.org), held in the NIMH Data Archive (NDA). This is a multisite, longitudinal study designed to recruit more than 10,000 children age 9-10 and follow them over 10 years into early adulthood. The ABCD Study® is supported by the National Institutes of Health and additional federal partners under award numbers U01DA041048, U01DA050989, U01DA051016, U01DA041022, U01DA051018, U01DA051037, U01DA050987, U01DA041174, U01DA041106, U01DA041117, U01DA041028, U01DA041134, U01DA050988, U01DA051039, U01DA041156, U01DA041025, U01DA041120, U01DA051038, U01DA041148, U01DA041093, U01DA041089, U24DA041123, U24DA041147. A full list of supporters is available at https://abcdstudy.org/federal-partners.html. A listing of participating sites and a complete listing of the study investigators can be found at https://abcdstudy.org/consortium_members/. ABCD consortium investigators designed and implemented the study and/or provided data but did not necessarily participate in the analysis or writing of this report. This manuscript reflects the views of the authors and may not reflect the opinions or views of the NIH or ABCD consortium investigators. The ABCD data repository grows and changes over time. The ABCD data used in this report came from <u>10.15154/z563-</u> <u>zd24</u>). DOIs can be found at https://nda.nih.gov/abcd/abcd-annual-releases.

## Author Contributions

Conceptualization, N.dL.; Methodology, N.dL., M.R., software, WL.; formal analysis, NdL., WL,; validation, N.dL., W.L, investigation, N.dL.; writing – original draft, N.dL., W.L., M.R.; writing – review & editing, N.dL., W.L., M.R., funding acquisition, N.dL.; resources, N.dL.; supervision, N.dL.

## Supplementary materials

**Supplementary Figure 1:**
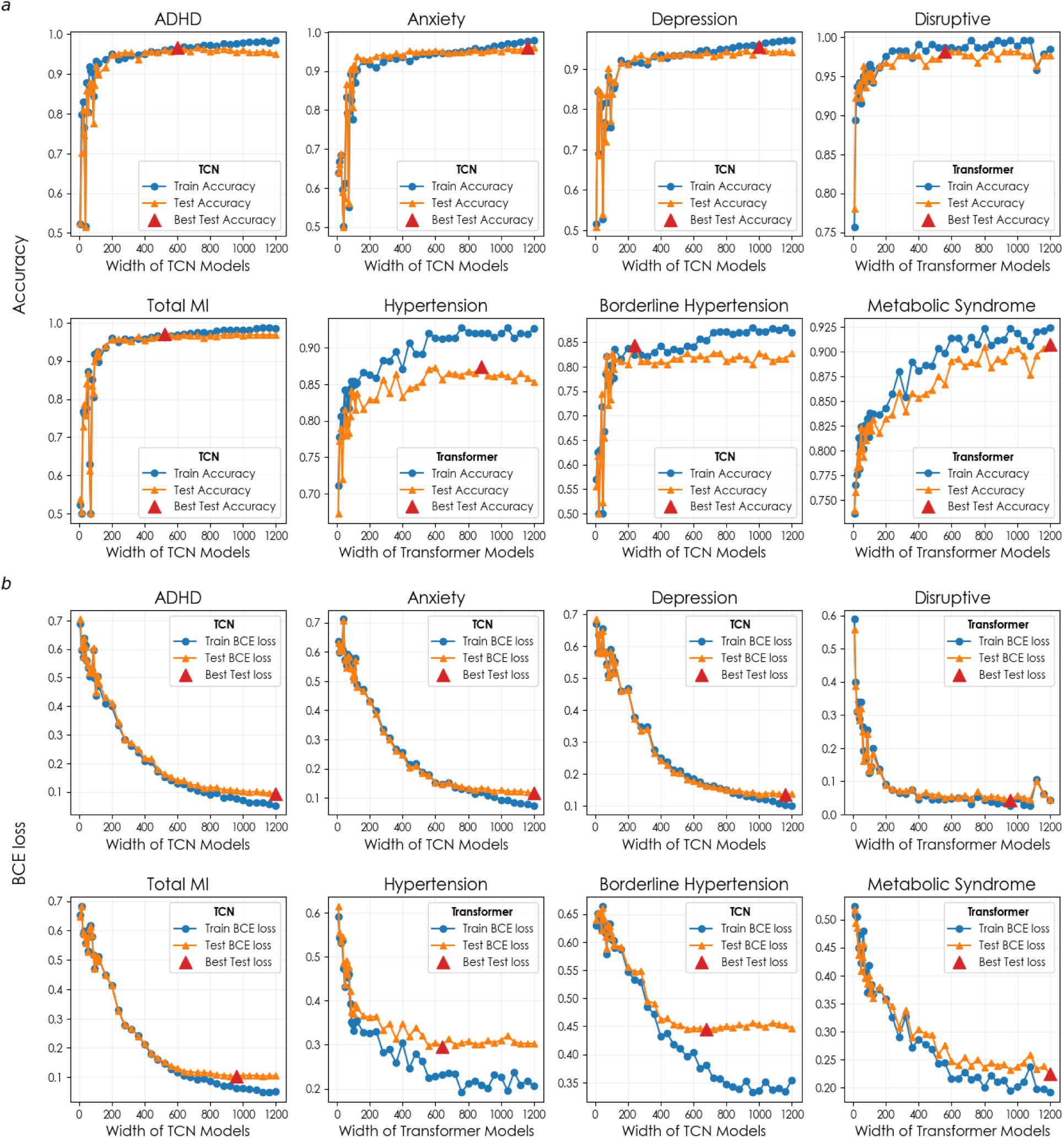
Performance-complexity tradeoffs in prediction. The relationship between increasing network width and accuracy (*a*) or binary cross-entropy loss (*b*) is shown for the best-performing timeseries model in out of sample for each predictive target. Red triangles indicate the optimal structural configuration identified by RiskPath.

**Supplementary Figure 2:**
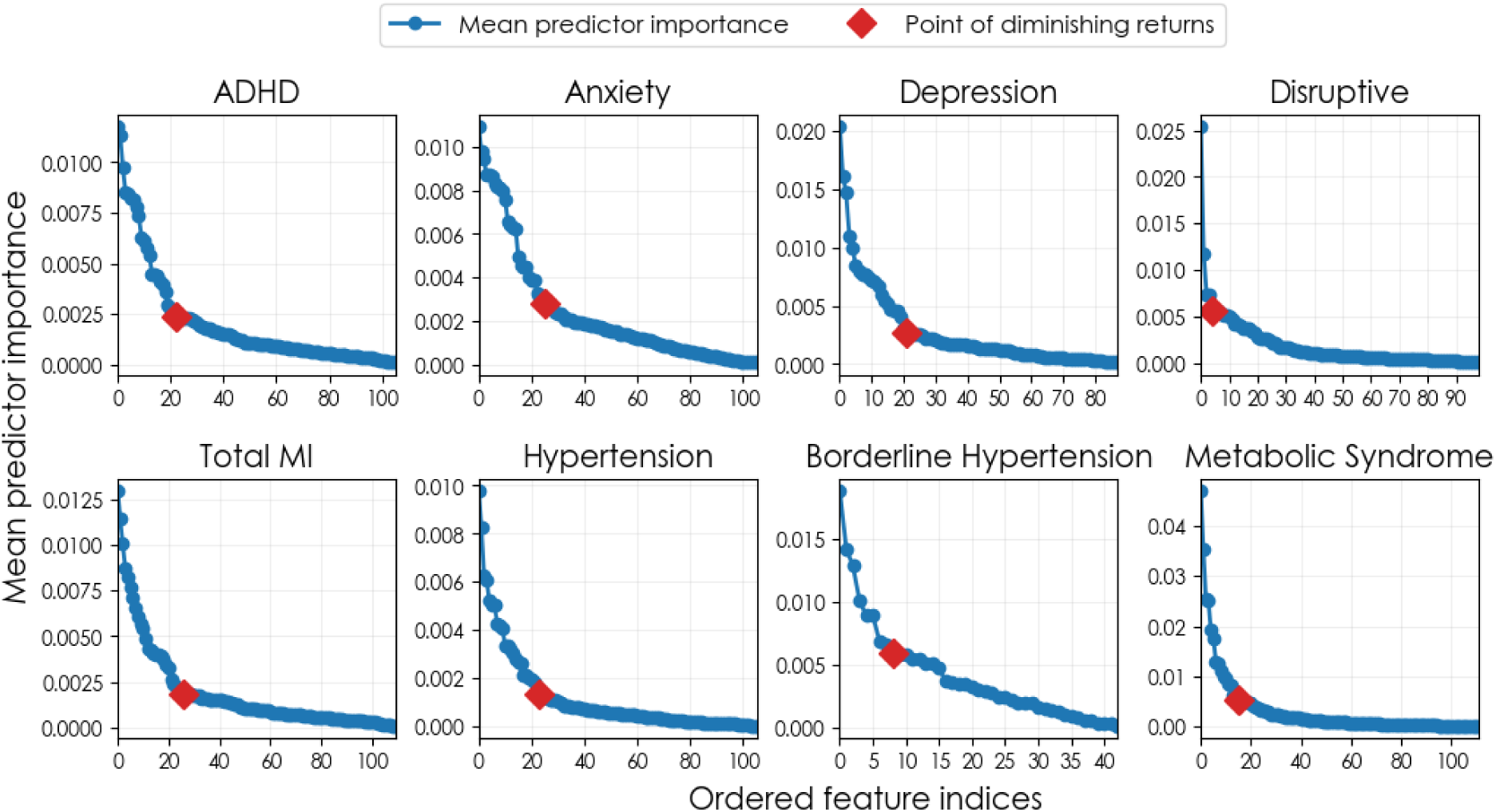
Feature ablation. The mean predictor importance of each predictive target for the best-performing timeseries model in out of sample testing is plotted to identify the point of diminishing returns (red).

**Supplementary Figure 3:**
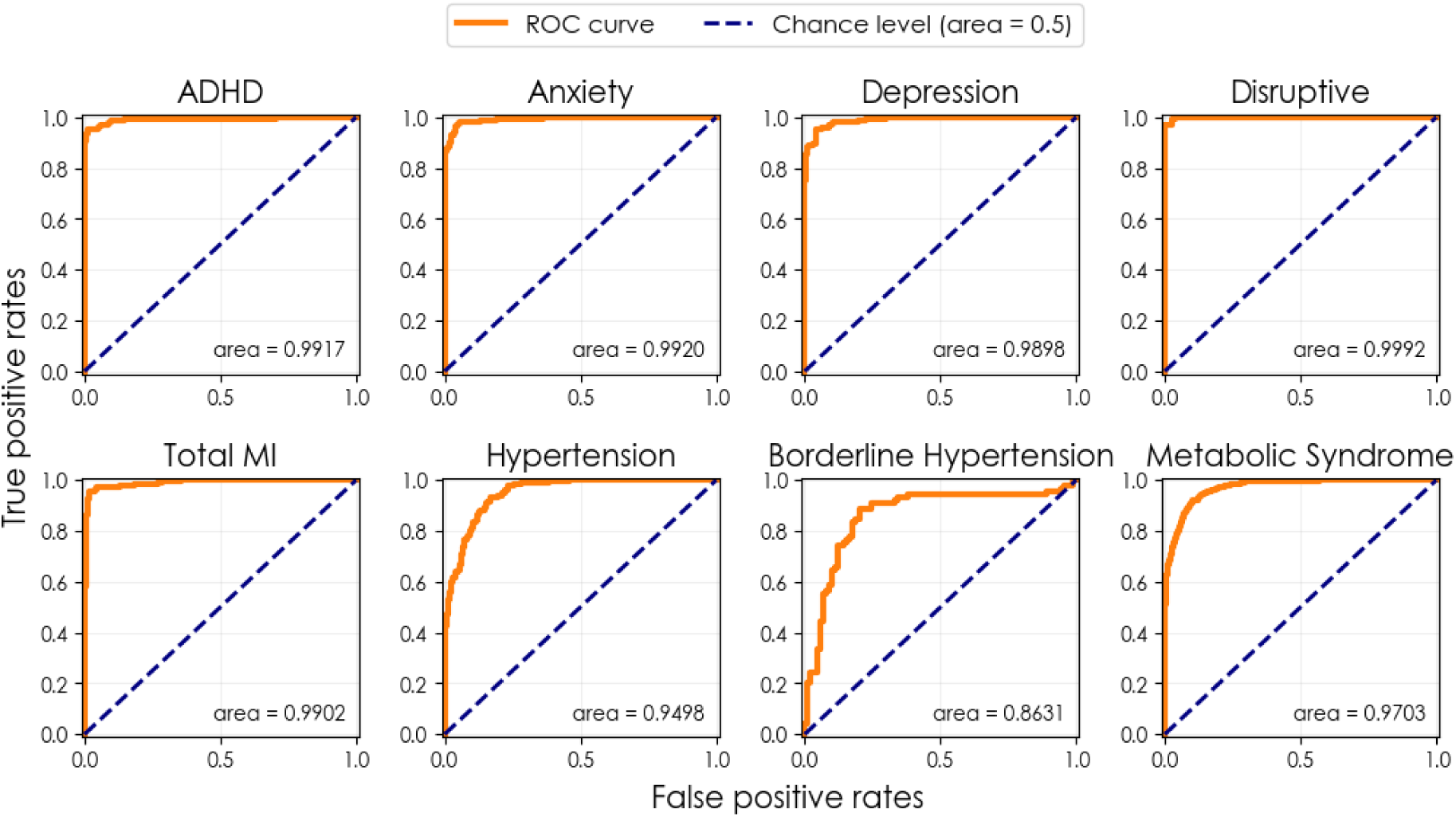
AUROC curves. The Area Under the Receiver Operating Characteristic (AUROC) curve of each predictive target with the best-performing timeseries model in out of sample testing.

**Supplementary Figure 4:**
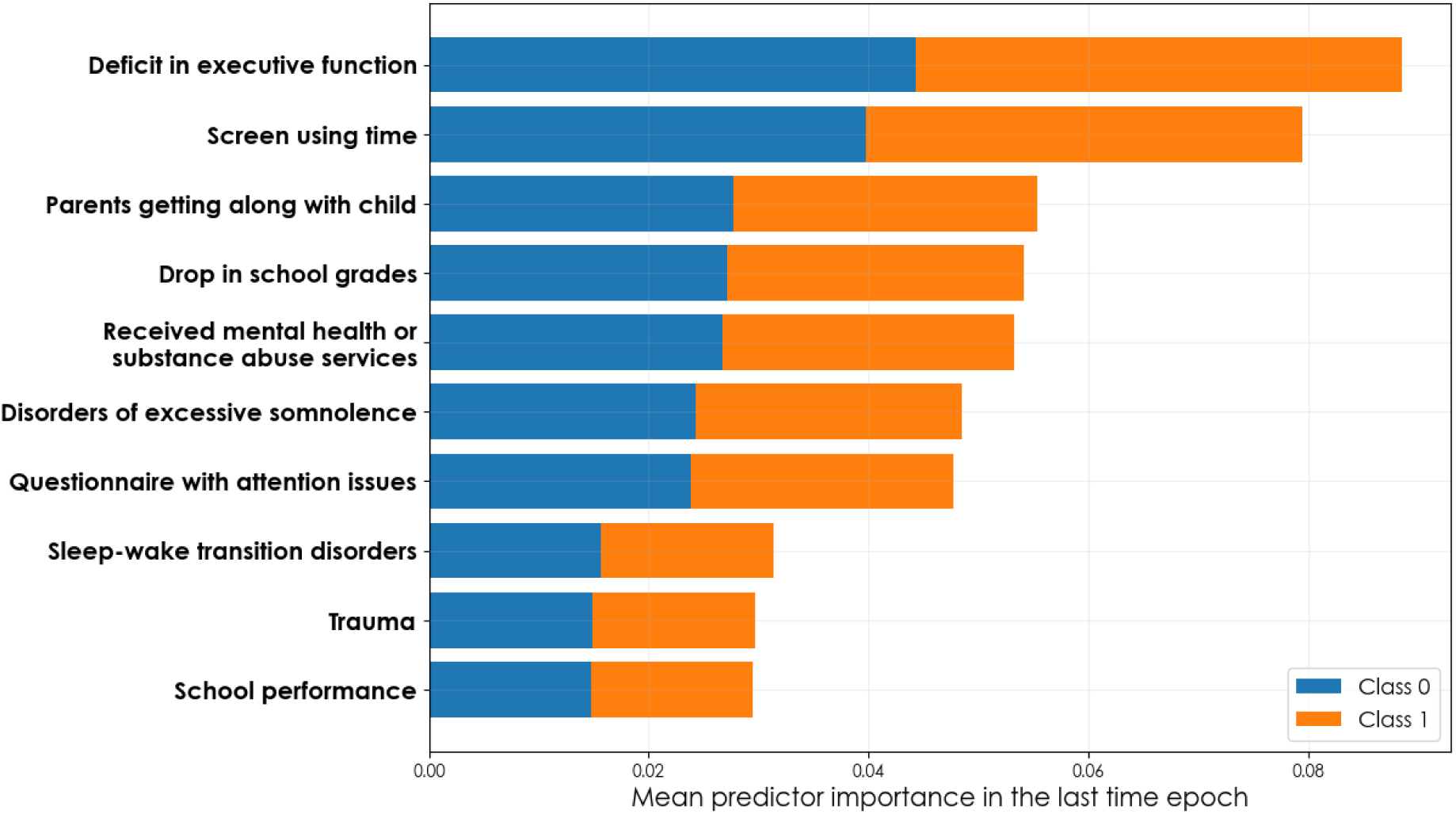
Visualization of mean predictor importance with a stacked bar chart. The mean predictor importances in the last time epoch (year) for predicting ADHD using the ablated Transformer model with 10 features. Different colors of each horizontal bar represent the mean predictor importance of the respective class of the binary target.

**Supplementary Figure 5:**
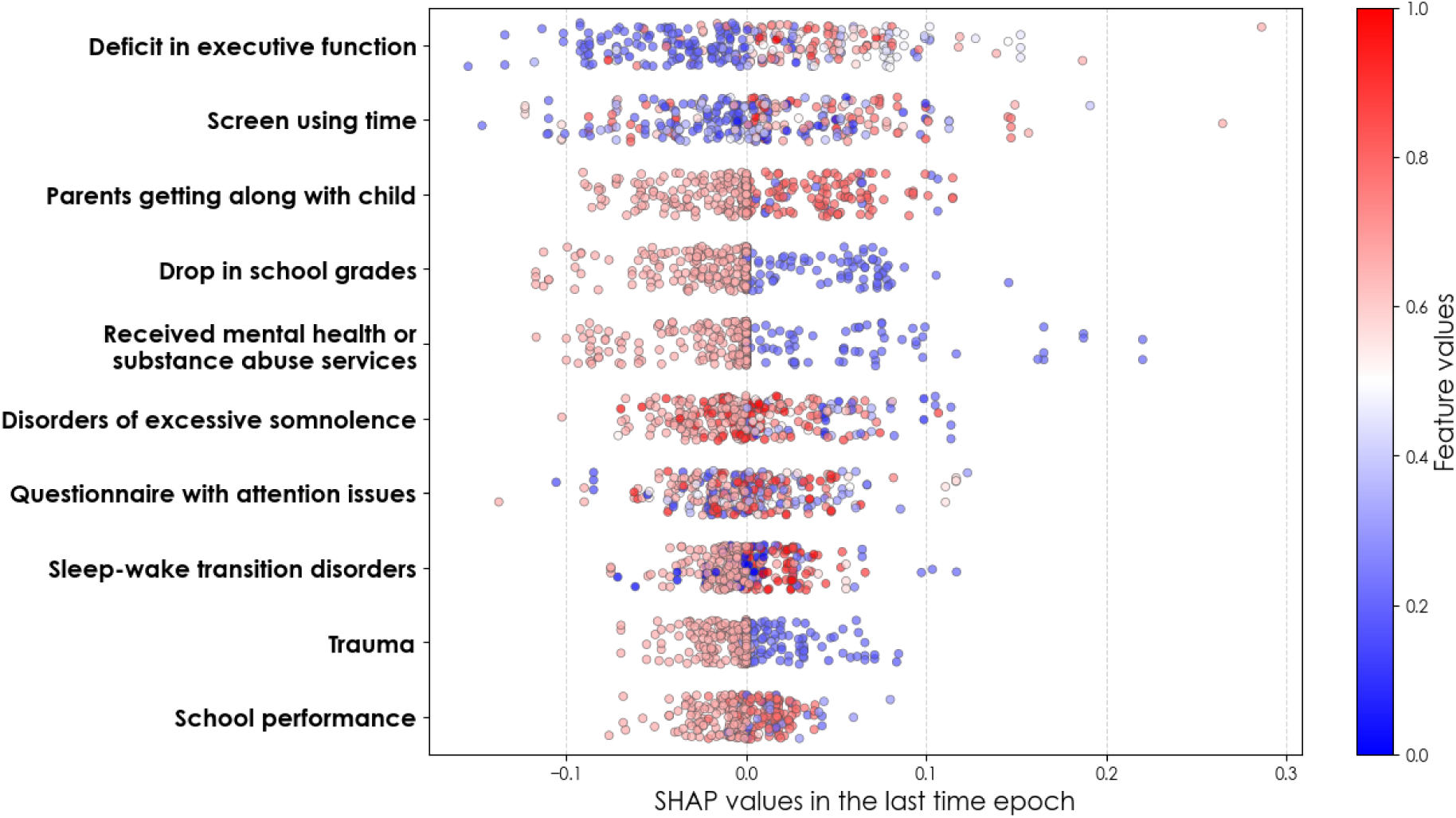
Visualization of SHAP values with a beeswarm plot. The SHAP values in the last time epoch (year) for predicting ADHD using the ablated Transformer model with 10 features. The colors of each data point encode the feature values (scaled to the unit interval).

**Supplementary Figure 6:**
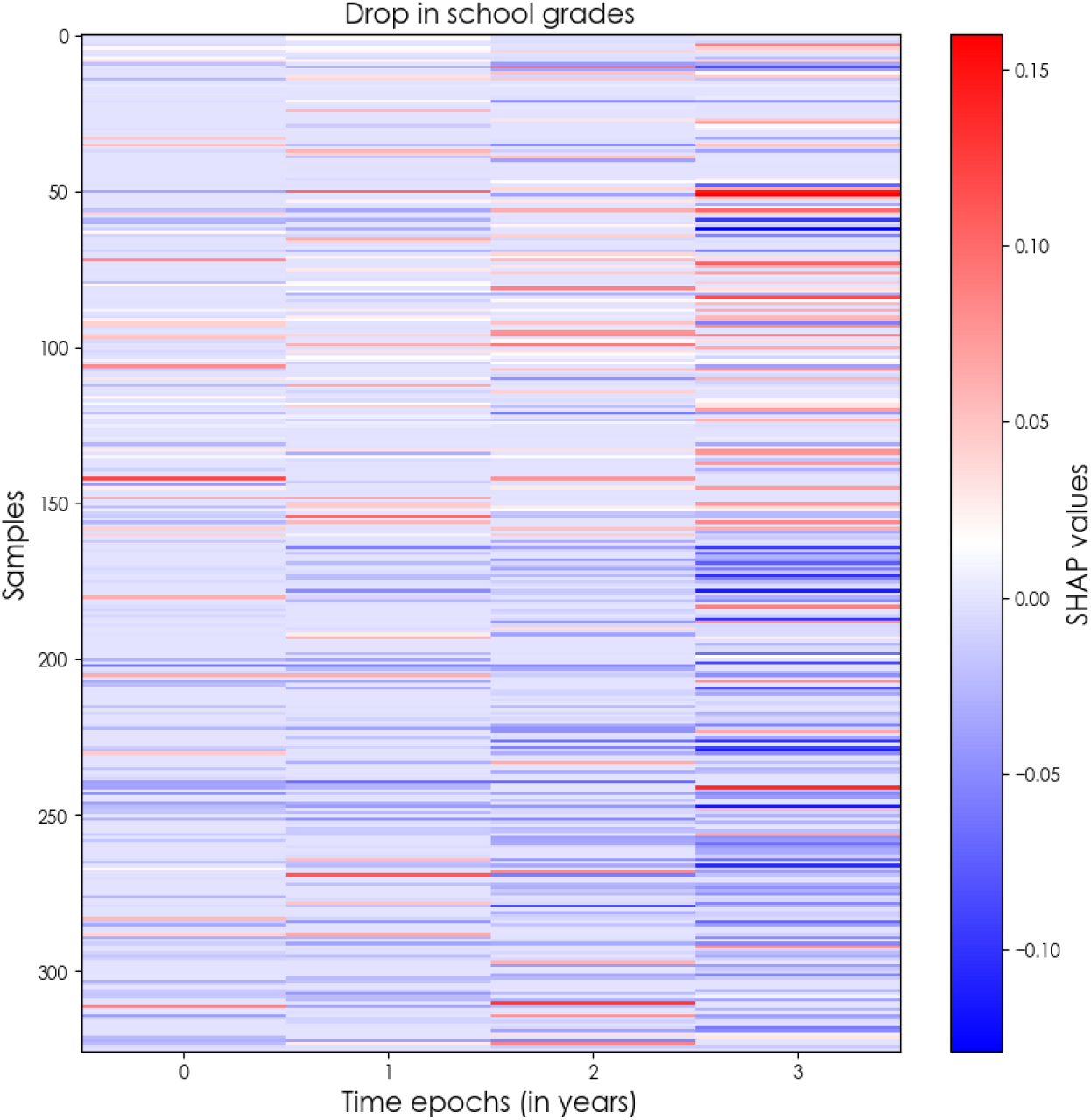
Visualization of SHAP values with a heatmap. The SHAP values of the binary feature encoding children’s drop in school grades for predicting ADHD using the ablated Transformer model with 10 features.

**Supplementary Table 1:**
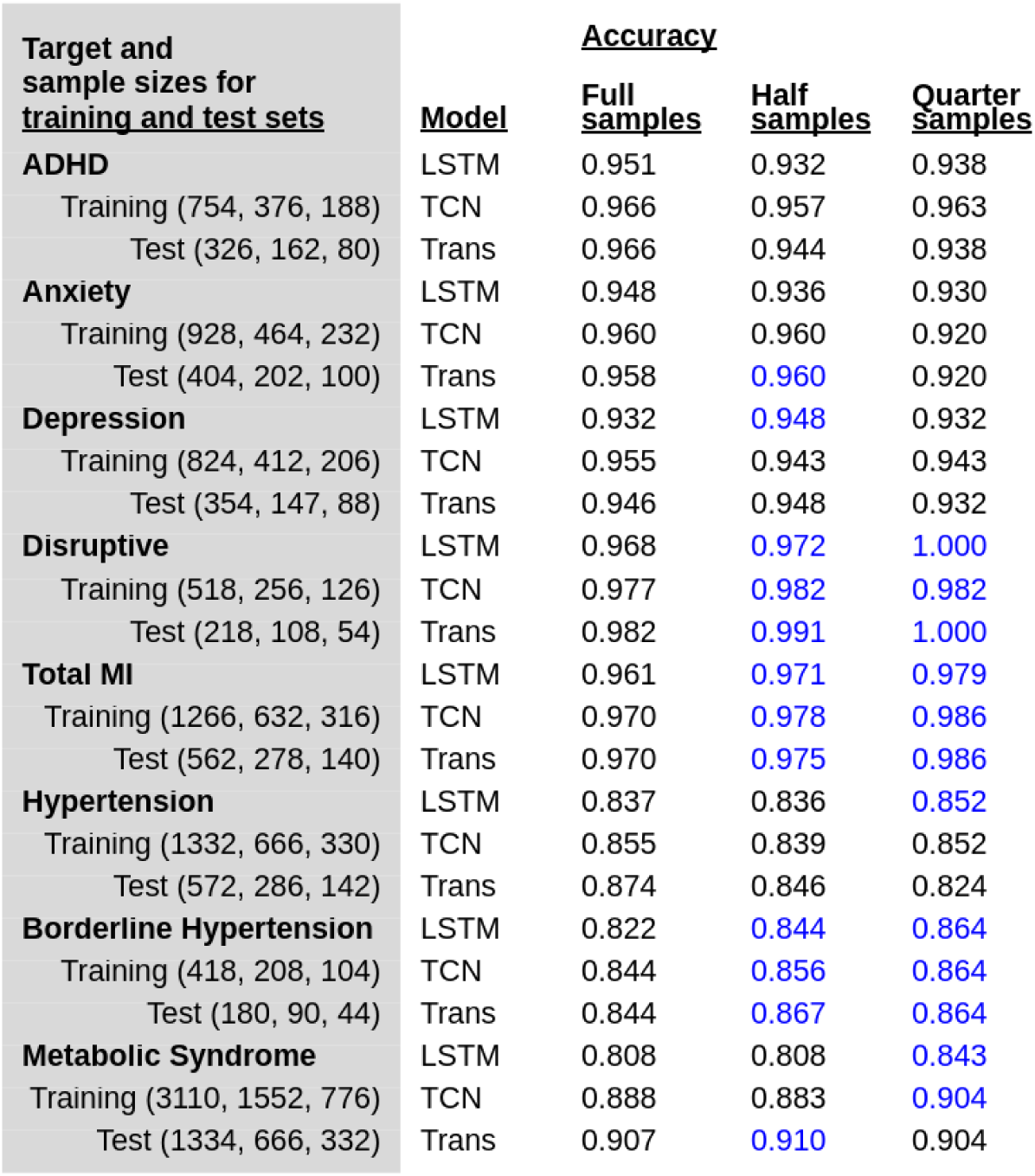
Sensitivity analysis under subsampling. Improvements in accuracy were observed after subsetting the full list of samples by 50% and 25% respectively, verifying the findings of Nakkiran et al.^94^ that a larger sample size can adversely affect model performance.

**Supplementary Table 3:**
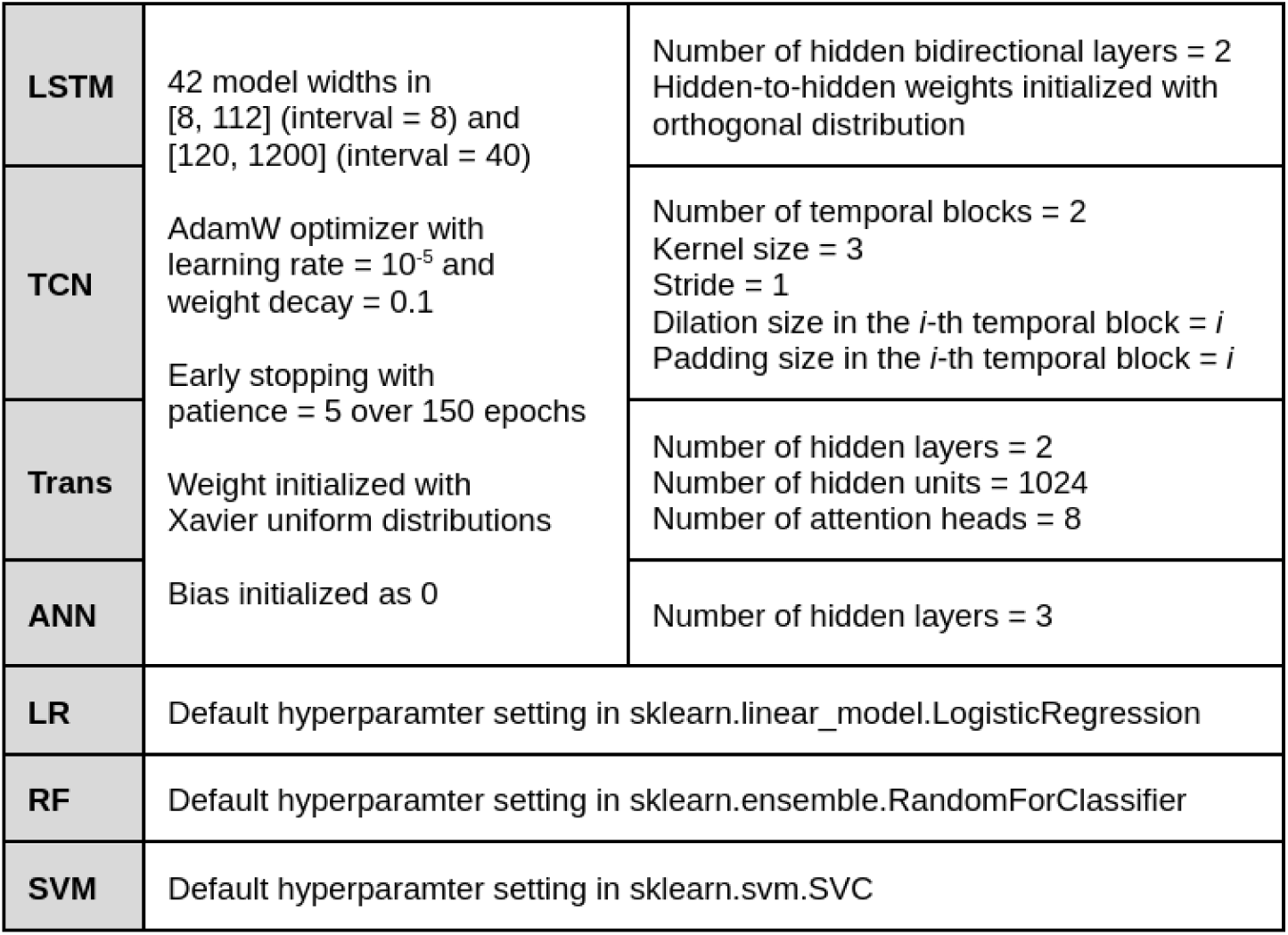
Algorithm setting and hyperparameters.

